# The *LRRK2* p.A419V variant associates with risk of Parkinson’s disease in the East Asian population and an evaluation on age of onset

**DOI:** 10.1101/2025.08.28.25333987

**Authors:** Kai Shi Lim, Maria Teresa Periñan, Elaine Guo Yan Chew, Paul Suhwan Lee, Fulya Akçimen, Jia Lun Lim, Mathew J Koretsky, Manabu Funayama, Hiroyo Yoshino, Nobutaka Hattori, Rauan Kaiyrzhanov, Henry Houlden, Mariam Isayan, Yi Wen Tay, Tzi Shin Toh, Lei-Cheng Lit, Anis Nadhirah Khairul Anuar, Hans Xing Ding, Laurel Screven, Norlinah Mohamed Ibrahim, Chin-Hsien Lin, Han-Joon Kim, Jee-Young Lee, Sun Ju Chung, Jia Nee Foo, Eng-King Tan, Shen-Yang Lim, Ai Huey Tan, Sara Bandres-Ciga, Azlina Ahmad-Annuar, the Global Parkinson’s Genetics Program (GP2)

**Affiliations:** Department of Biomedical Science, Faculty of Medicine, Universiti Malaya, 50603 Kuala Lumpur, Malaysia; Department of Medicine, Faculty of Medicine, Universiti Malaya, 50603 Kuala Lumpur, Malaysia; Unidad de Trastornos del Movimiento, Servicio de Neurología y Neurofisiología Clínica, Instituto de Biomedicina de Sevilla, Hospital Universitario Virgen del Rocío, Consejo Superior de Investigaciones Científicas (CSIC), Universidad de Sevilla, Seville, Spain; Centre for Preventive Neurology, Wolfson Institute of Population Health, Queen Mary University of London, London, United Kingdom; Lee Kong Chian School of Medicine, Nanyang Technological University Singapore, Singapore, Singapore; Center for Alzheimer’s and Related Dementias, National Institute on Aging and National Institute of Neurological Disorders and Stroke, National Institutes of Health, Bethesda, Maryland, USA; Laboratory of Neurogenetics, National Institute on Aging, National Institutes of Health, Bethesda, MD, USA; The Mah Pooi Soo & Tan Chin Nam Centre for Parkinson’s & Related Disorders, Faculty of Medicine, Universiti Malaya, Kuala Lumpur, Malaysia; Department of Neurology, Faculty of Medicine, Juntendo University, 2-1-1 Hongo, Bunkyo-ku, Tokyo, 113-8421, Japan; Research Institute for Diseases of Old Age, Graduate School of Medicine, Juntendo University, 2-1-1 Hongo, Bunkyo-ku, Tokyo, 113-8421, Japan; International Collaborative Research Administration, Juntendo University, Tokyo, Japan; Department of Neuromuscular Diseases, UCL Queen Square Institute of Neurology, University College London, London, UK; South Kazakhstan Medical Academy, Department of Neurology, 1/1 Al-Farabi Avenue, 160019 Shymkent, Kazakhstan; Department of Neurology and Neurosurgery, National Institute of Health, Yerevan, Armenia; Department of Physiology, Faculty of Medicine, Universiti Malaya, 50603 Kuala Lumpur, Malaysia; The Global Parkinson’s Genetics Program (GP2); Department of Medicine, Faculty of Medicine, Universiti Kebangsaan Malaysia, Selangor, Malaysia; Department of Neurology, National Taiwan University Hospital Taipei, Taipei, Taiwan; Department of Neurology, Seoul National University Hospital, College of Medicine, Seoul National University, Seoul, Republic of Korea; Department of Neurology, Seoul Metropolitan Government - Seoul National University Boramae Medical Center, Seoul National University College of Medicine, Seoul, Korea; Department of Neurology, Asan Medical Center, University of Ulsan College of Medicine, Seoul, South Korea; Duke-National University of Singapore Medical School, Singapore, Singapore; Department of Neurology, National Neuroscience Institute, Singapore General Hospital, Singapore, Singapore

**Keywords:** Parkinson’s disease, genetics, *LRRK2*, A419V, risk, age at onset, Asian

## Abstract

Common and rare variants in *LRRK2* influence Parkinson’s disease (PD) risk across diverse populations. We investigated the p.A419V variant across multiple ancestry cohorts comprising over 200,000 individuals. In cases of East Asian ancestry, the variant was significantly associated with increased risk (OR = 2.9; 95% CI: 1.66–5.10; p = 0.0002), It lies on rare EAS haplotypes, and was not in linkage disequilibrium with other *LRRK2* coding variants. Although not significant, meta-analysis of age of onset in EAS cases show a suggestive trend (β = –0.89 years; SE = 1.01; *p* = 0.380). LRRK2 protein modelling prediction indicated that binding sites for RAB8A, RAB10, RAB29 were in close proximity to the p.A419V variant within the ARM domain. Together, these findings confirm the p.A419V as a significant PD risk factor in EAS populations, as well as highlight disease-relevant variants in the ARM domain and the link with LRRK2-RAB signaling pathway.

## Introduction

Pathogenic variants in the LRRK2 gene, including p.R1067Q, p.N1437H, p.R1441G/C/H, p.Y1699C, p.G2019S and p.I2020T, are known to cause Mendelian forms of Parkinson’s disease (PD), and several were also identified as risk variants through genome-wide association studies (GWAS) and case-control analyses^1–4^. LRRK2 ‘Asian risk variants’, p.R1628P and p.G2385R, have been identified as key risk factors for sporadic PD in various Asian populations^5,6^.

Another LRRK2 variant, p.A419V (chr12:40252984:C>T, rs34594498, NM_198578.4:c.1256C>T), was first studied as a potential PD-associated variant in East Asians (EAS) from Taiwan^7^, where no significant association was found in a cohort of 608 cases and 373 controls. In a subsequent, larger combined EAS cohort from Japan, Korea, and Taiwan (1,376 cases; 962 controls), p.A419V was reported as a significant risk variant (odds ratio [OR] = 2.7; 95% confidence interval [CI] = 1.35 - 3.83; p = 0.0045)^1^. Several studies in EAS populations attempting to replicate this finding reported varied and inconsistent results^8–12^. In the most recent PD GWAS in EAS from mainland China, this variant reached suggestive genome-wide significance in both the discovery and replication cohorts (discovery: n = 1,972 cases, 2,478 controls; p = 2.47e-06, OR = 2.0; 95% CI = 1.33 - 3.03; replication: n = 8,209 cases, 9,454 controls; p = 2.64e-15, OR = 2.4; 95% CI = 1.90 - 2.90)^13^. EAS cohorts from 23andMe included in a multi-ancestry meta-analysis^14^ also showed a significant association between this variant and PD risk (n = 322 cases, 151,905 controls; OR = 3.2; 95% CI = 1.85 - 5.61; p = 0.0003).

In the broader Asian context, the p.A419V variant was not reported as significant in a case-control study from Kazakhstan in Central Asia (minor allele frequency, MAF n = 292 cases, 199 controls; OR = 1.5, p = 0.4)^15^. In South Asian cohorts, the p.A419V variant is rare with a MAF = 0.4% (n = 4,806 cases, 6,364 controls)^16^. The variant is absent in studies on Vietnamese (n = 83 early-onset PD (EOPD) cases^17^, and there is no available information from recent studies in Japan^18^ (n = 221 cases), Thailand^19^ (n = 47 EOPD cases), or Korea^20^ (GWAS n = 1,050 cases, 5,000 controls). This variant is absent/rare in European^14,21^ (n = 37,688 cases, 18,618 proxy-cases, 1.4 million controls; n = 49,049 cases, 18,785 proxy cases and 2,458,063 controls), African^22^ (GWAS n = 1,488 cases; 196,430 controls), Latin American^23^ (1,734 cases and 1,097 controls; Loesch et al., 2021, n = 807 cases and 690 controls), and North African Egyptian^24^ PD cohorts. These differences in LRRK2 p.A419V detection may be due to the higher allele frequency of p.A419V in EAS (MAF = 0.01028) compared to MAF < 0.001 in European, South Asian, and Middle East populations, and an even lower frequency in African populations (MAF < 0.0001), as reported in gnomAD v4.1.0 (exome + genome).

LRRK2 activity assays indicate that the p.A419V variant increases LRRK2-mediated Rab10Thr73 phosphorylation by more than 1.5-fold compared to wild-type, and moderately elevates the formation of LRRK2 filaments in the absence of MLi-2 treatment, consistent with other pathogenic LRRK2 variants^25^.

Despite evidence pointing to a potential deleterious effect of the LRRK2 p.A419V variant, inconsistent findings of variable significance across genetic association studies limited its inclusion in further functional characterization and clinico-genetic correlation efforts, particularly in comparison to the well-established EAS LRRK2 variants such as p.G2385R and p.R1628P^26^. The relevance of this variant in other PD populations was not fully studied. To address this gap while also increasing confidence for deep phenotyping and precision medicine applications, we leveraged large-scale genotyping data from the Global Parkinson’s Genetics Program (GP2), whole-genome sequencing (WGS) data from the Accelerating Medicines Partnership Parkinson’s Disease (AMP-PD)^27,28^, the UK Biobank (UKB, www.ukbiobank.ac.uk), and the All of Us Research Program (AOU, https://allofus.nih.gov/) in addition to whole-exome sequencing (WES) data from an EAS dataset^29^ and Juntendo University Hospital PD cohort. In addition, there is inconclusive evidence of the influence of Asian LRRK2 risk variants on age of onset^30,31^; therefore, this study sought to address this further for the p.A419V variant.

## Methods

### Cohorts under study

This study included six cohorts spanning multiple ancestries. Cohort 1 included genotyped data from GP2 release 9 (https://gp2.org/), comprising 25,699 unrelated PD patients and 13,652 controls from ten ancestry populations: European (EUR), East Asian (EAS), Admixed American/Latin American (AMR), Ashkenazi Jews (AJ), Central Asian (CAS), Complex Admixture History (CAH), Middle Eastern (MDE), South Asian (SAS), African American (AAC) and African (AFR) (Supplementary Table 1). Cohort 2 was composed of data from AMP-PD WGS release 3 (https://amp-pd.org/), which included 2,251 unrelated PD cases and 2,835 controls of European descent (Supplementary Table 1). Cohort 3 comprised individuals from the UKB (2,954 cases and 56,256 controls). Cohort 4 included data from the AOU Program (2,103 cases, 29,733 controls). Cohort 5 was an EAS-WES replication cohort (3,967 cases, 5,457 controls) described further below. Cohort 6 was a separate EAS cohort of Japanese ancestry (2,729 PD cases recruited from Juntendo University Hospital and 61,332 control individuals from the public database jMorp 61KJPN^32^, https://jmorp.megabank.tohoku.ac.jp/). Genotype comparisons in the Japanese population were conducted using R software, including a two-tailed Fisher’s exact test for calculation of OR for allele frequencies, and 95% CI.

### Data Quality Control

Quality control (QC) of the GP2 data was performed using the GenoTools pipeline (https://github.com/GP2code/GenoTools)^33^. Briefly, samples were excluded if they had a genotyping rate below 95%, exhibited sex mismatches, or were duplicated (KINSHIP > 0.354), or displayed high heterozygosity (|F| statistic > 0.25). Variants were excluded if they had > 5% missingness, significant deviations from Hardy-Weinberg Equilibrium (HWE p < 1e-4), or non-random missingness based on case-control status (p ≤ 1e-4). Ancestry estimation was performed using Genotools^33^, with the default reference panels from the 1000 Genomes Project, the Human Genome Diversity Project, and Ashkenazi Jewish datasets. Additionally, individuals with second-degree or closer relatedness (KINSHIP > 0.0884) were removed before analysis. The percentage of ancestry was then calculated using the supervised functionality of ADMIXTURE (v1.3.0; https://dalexander.github.io/admixture/binaries/admixture_linux-1.3.0.tar.gz)^34^ with the same reference panel mentioned before to estimate the ancestry proportions of the GP2 data accurately.

### Statistical analyses

The LRRK2 p.A419V variant was extracted from the raw genotyped NeuroBooster Array v1.0 (NBA)^35^ (Supplementary table 1) for GP2 data release 9 and from WGS data for AMP-PD version 3, the UKB, and the AOU.

LRRK2 p.A419V PD carriers and non-carriers were compared for sex and family history using the two-tailed Fisher’s exact test. Association between the LRRK2 p.A419V variant and PD risk was assessed by performing a logistic regression (glm) under an additive genetic model in PLINK 2.0 (Chang et al. 2015), where genotypes were encoded as 0 (homozygous major allele), 1 (heterozygous), and 2 (homozygous minor allele). Covariates, including sex, and the appropriate number of principal components (PCs) were included in the model. The number of PCs was determined by identifying the elbow point of the scree plot (Supplementary Figure 2) to account for population stratification (Supplementary Table 2)^36^. Additionally, allele and genotype frequencies, as well as HWE, were calculated using PLINK 2.0. Power calculation was performed using the GAS power calculator (https://csg.sph.umich.edu/abecasis/cats/gas_power_calculator/). The additive model was used with a significance level set at p = 0.05, PD general population prevalence of 0.5%^21^ at an OR of 2.27 according to the meta-analysis of this variant in the Asian population^1^ and an OR of 2.01 according to the largest Chinese GWAS study^13^. The association between p.A419V and age at onset (AAO) was assessed using a linear regression additive genetic model adjusted for sex and the appropriate number of PCs, as described above. Meta-analysis of AAO between GP2 EAS discovery cohort and EAS-WES replication cohort was analysed using R meta package^37^.

The EAS replication cohort consisted of published WES data from 3,967 PD patients and 5,457 ancestry- and geographically matched controls from five regions across East Asia [Singapore (SG): 1,955 cases, 3,630 controls; Malaysia (MAL): 325 cases, 59 controls; Hong Kong (HK): 70 cases, 586 controls; South Korea (KR): 1,417 cases, 1,040 controls; Taiwan (TW): 200 cases, 142 controls] ^29^. A stratified Cochran-Mantel-Haenszel (CMH) test was used to evaluate the burden of LRRK2 p.A419V across the exomes of participants from the replication cohort. Fisher’s exact test (two-tailed) was used to assess the burden of LRRK2 p.A419V within each stratum. SG and MAL samples were considered as one stratum due to the similarity in genetic background.

Cohort 6 was a second EAS replication cohort of Japanese ancestry (2,729 PD cases recruited from Juntendo University Hospital with age of onset ≥ 21 and 61,000 control individuals from the public database jMorp 61KJPN^32^, https://jmorp.megabank.tohoku.ac.jp). Genotype comparisons in the Japanese population were conducted using R software, including a two-tailed Fisher’s exact test for calculation of odds ratios (OR) for allele frequencies, and 95% confidence intervals (CI).

Haplotype blocks in the GP2 EAS cohort were defined using the --block function in PLINK 1.9, with the minimum MAF threshold set to 0.0001 to allow the inclusion of LRRK2 p.A419V across all ancestries. Haplotype frequency estimation and association analysis were performed using the haplo.stats R package (version 1.9.7; https://cran.r-project.org/web/packages/haplo.stats/) under default settings. To further characterize the linkage disequilibrium (LD) structure, pairwise r² values between LRRK2 p.A419V and all LRRK2 missense variants and other reported GWAS variants in the LRRK2 locus^1–4,14,38,39^ were calculated using PLINK 1.9.

### LRRK2 protein structure prediction

The three-dimensional structures of LRRK2, and complexes with RAB8A, RAB10, RAB29, RAB32 were predicted using AlphaFold3 (https://hpc.nih.gov/apps/alphafold3/)^40^ on the NIH Biowulf high-performance computing cluster (https://hpc.nih.gov). FASTA sequences for each protein (LRRK2:Q5S007, RAB8A:P61006, RAB10:P61026, RAB29:O14966, RAB32:Q13637) were retrieved from UniProt (https://www.uniprot.org/)^41^, and subsequently AlphaFold3 input JSON files were generated. Multiple sequence alignments and model inference were performed to generate predicted protein structures. To improve the robustness of the predictions, five independent runs were initiated using different random seeds, and then the model with the best ranking score for each complex was selected for visualization using the PyMOL Molecular Graphics System, Version 3.0 Schrödinger, LLC.

## Results

### Overall data review

Demographics for each cohort separated by ancestry, including age at sample collection, age at onset, sex, and number of cases/controls, are presented in Supplementary Table 1. The LRRK2 p.A419V variant was in HWE across all ancestry groups in all the cohorts (Supplementary Table 4).

In the GP2 cohort, the LRRK2 p.A419V variant was found to be rare (MAF < 1%) across all populations (Table 1), with the highest frequency observed in the EAS (MAF = 1.3% cases, 0.36% controls), followed by CAS (MAF = 0.98% cases, 0.6% controls), CAH (MAF = 1.1% cases, none in controls), EUR (MAF = 0.12% cases, 0.02% controls), AJ (MAF = 0.03% cases, none in controls). In contrast, no carriers were observed in the AAC, AFR, AMR, MDE and SAS individuals. None of the cases were carriers in the EUR AMP-PD cohort, while the MAF in controls was 0.02%. In the UKB cohort, no carriers were identified in AAC, AFR, AMR, CAH, CAS, FIN, or MDE ancestry groups, while the MAF in EAS controls was 0.65% and no carriers were identified in the six EAS cases. The variant was present in UKB EUR individuals with MAF = 0.02% (cases) and MAF = 0.01% (controls). The MAF in the UKB SAS was 1.1% cases and 0.08% controls. In the AOU cohort, the highest number of carriers were of EAS (MAF = 6.60% cases, 0.40% controls), followed by EUR (MAF = 0.03% cases, 0.03% controls) and AMR (MAF = 0.01% controls, none in cases) ancestries. Thus, taking shared ancestries across the GP2, AMP-PD, UKB, and AOU cohorts, the EAS (MAF = 0.91%), CAS (MAF = 0.71%), CAH (MAF = 0.62%), SAS (MAF = 0.07%), EUR (MAF = 0.03%), AJ (MAF = 0.02%) (Table 1).

**Table 1.**
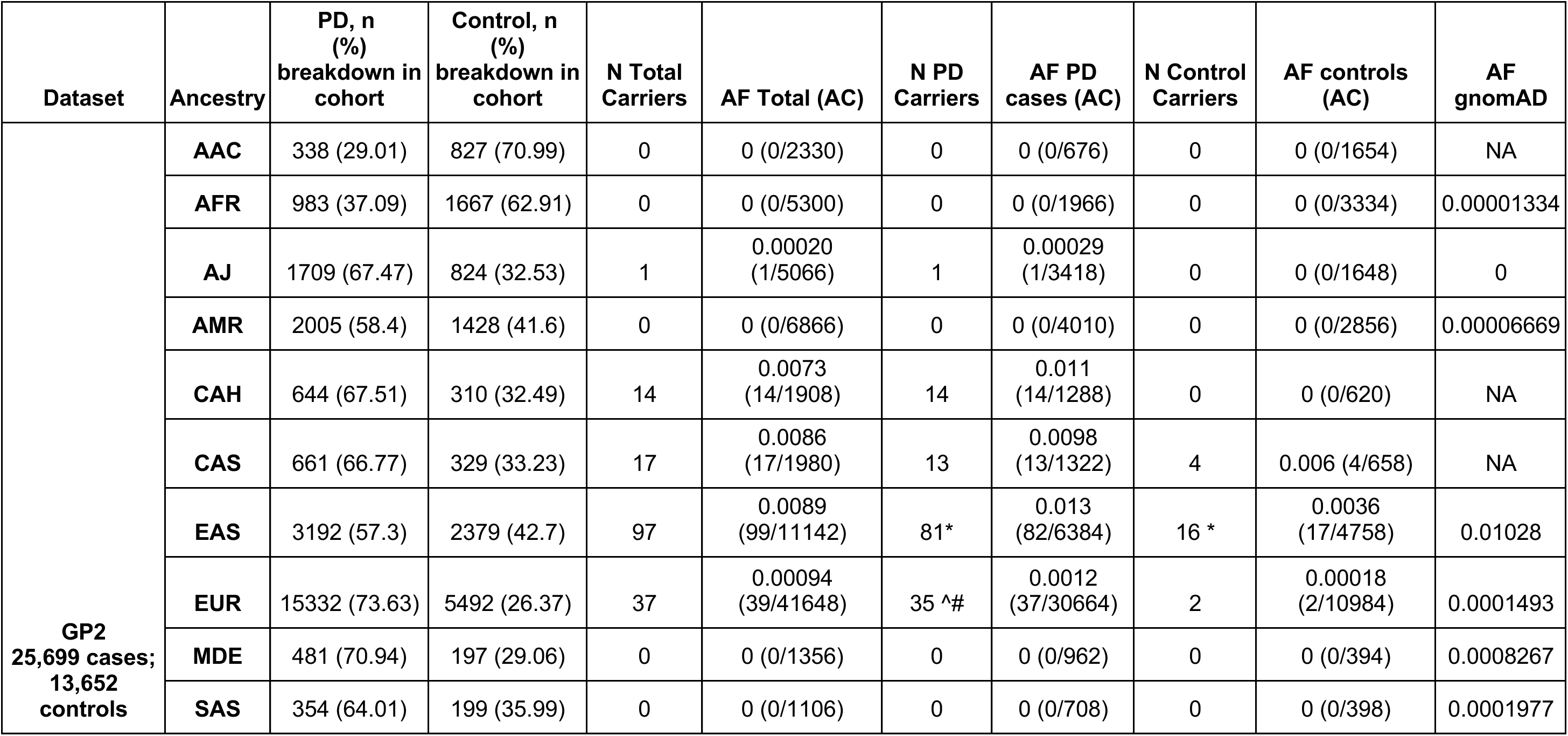
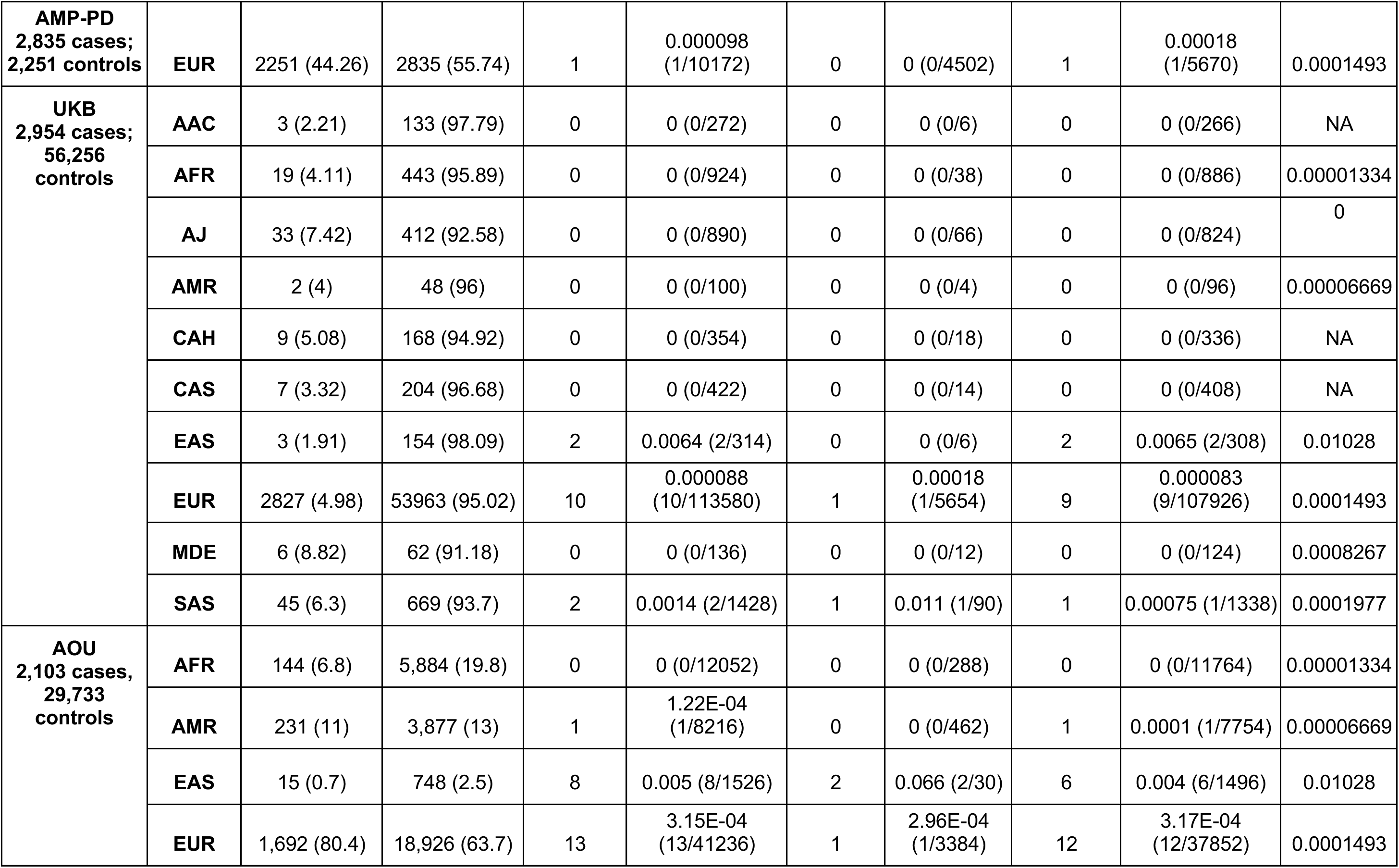
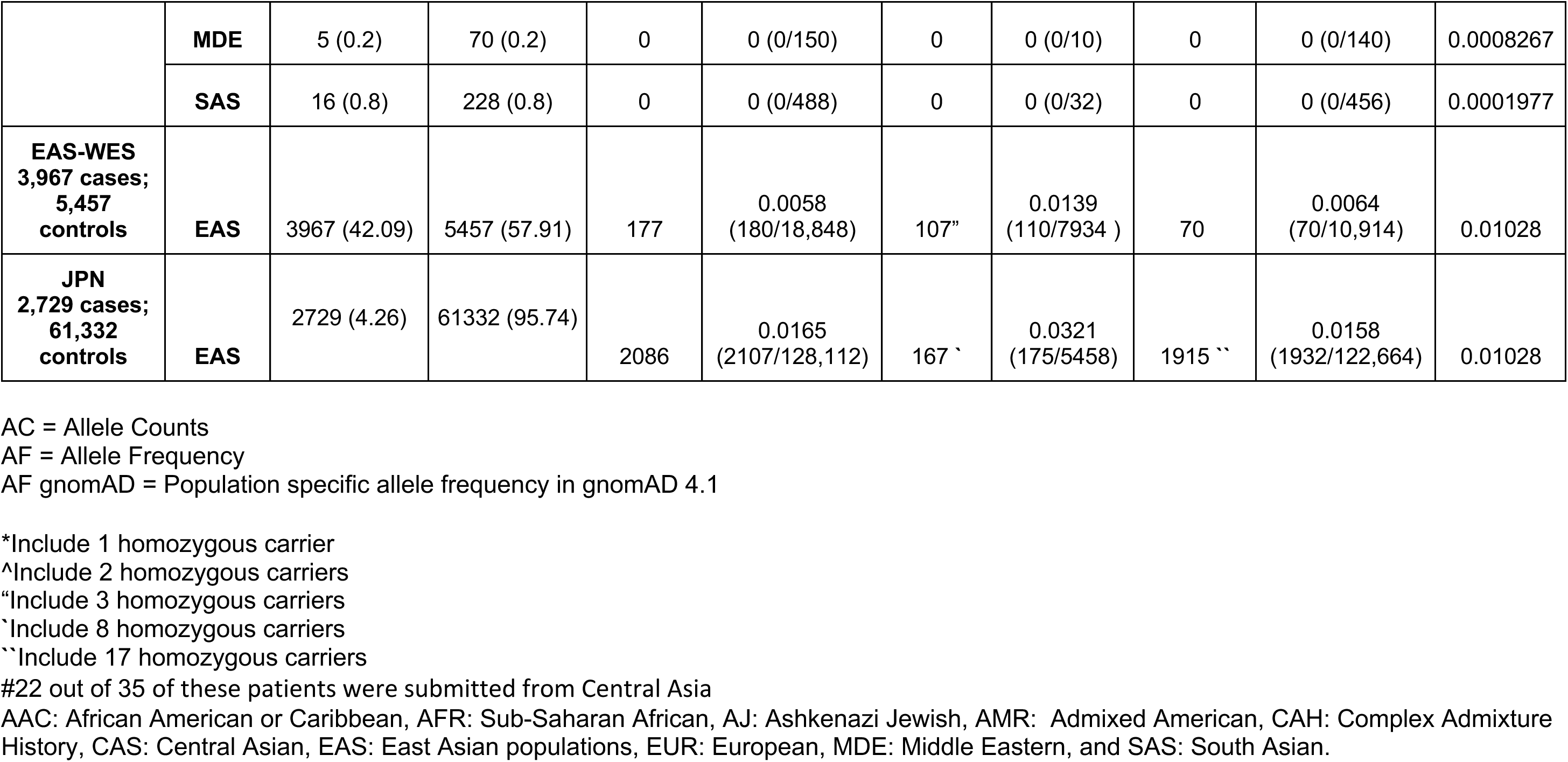
Frequency of the *LRRK2* p.A419V variant across ancestries.

Family history was not more commonly reported in LRRK2 p.A419V carriers (17.7%) compared to PD non-carriers across all ancestries, where it was reported in 25.2% of individuals (Table 1). A higher frequency of female LRRK2 p.A419V carriers was observed in the GP2 CAS and EUR groups (CAS: p = 0.008, 92.3% in carriers vs. 53.7% in non-carriers; EUR: p = 0.041, 54.6% in carriers vs. 37.7% in non-carriers) (Table 1).

### Risk association analysis

Power calculations indicated that only the EAS and EUR cohorts in GP2 had an 80% power to detect an association with an OR > 2.0 and a p-value < 0.05 (Supplementary Table 3), while the AMP-PD, UKB, and AOU cohorts were underpowered for association analysis of the LRRK2 p.A419V variant.

Logistic regression analyses conducted in the GP2 EAS and EUR ancestry groups indicated that a significantly higher frequency of the LRRK2 p.A419V variant was observed in cases compared to controls in both the EAS (MAF cases = 1.15% vs. MAF controls = 0.57%; OR = 2.908; 95% CI = 1.659 - 5.098, p = 0.0002) and EUR (MAF cases = 0.06% vs. MAF controls = 0.02%; OR = 5.754; 95% CI = 1.399 - 23.66, p = 0.015) groups (Supplementary Table 5). The association in the EAS group was tested in the independent EAS cohort from the EAS exome dataset^29^. The LRRK2 p.A419V variant was present in 107 of 3,967 cases (1.4%) and 70 of 5,457 controls (0.6%), with a significant risk association (OR = 1.51, 95% CI =1.103 - 2.068, p = 0.012, Supplementary Table 6). In the Juntendo EAS-Japanese replication cohort, the p.A419V variant was significantly associated as a risk factor in Japanese PD patients (MAF = 3.20%) than in the controls (MAF = 1.58%); (OR = 2.06; 95% CI: 1.76 - 2.42; p = 1.301 × 10⁻16, Supplementary Table 6).

In the GP2 cohort, there was minimal LD (r² < 0.01) between p.A419V and other LRRK2 coding variants across all ancestries studied (Supplementary Table 9), suggesting that the observed risk association is unlikely to be confounded by nearby coding variation. In addition, the LRRK2 p.A419V variant is not in LD with any lead SNPs identified in GWAS of PD in EAS populations^4,14,38,39^.

The LD block constructed around p.A419V consists of 8 SNPs (rs10506148, rs28365214, rs10878249, p.A211V, rs732374, p.V366M, p.L378F, rs1491938). Six rare haplotypes (MAF < 0.01) were identified in the GP2 EAS cohort, with one (Haplotype 9) observed in 1.2% PD cases and 0.34% controls (Supplementary Table 10), but further analysis lacked sufficient power to assess association. The haplotype analysis was not performed in the other EAS-WES dataset, as its exome sequencing data would not have fully contained sufficient variants to perform the LRRK2 haplotyping effectively. Additionally, the UKB and AOU cohorts had relatively small EAS sample sizes, limiting their utility for this analysis. No haplotype containing LRRK2 p.A419V was identified in the other GP2 ancestries due to the rarity of the variant.

### Admixture analysis of the GP2 EUR cohort

As the comparatively higher allele frequency and risk association of the LRRK2 p.A419V in European individuals was unique to the GP2 EUR cohort - unlike the other EUR cohorts from AMP-PD, UKB, and AOU - carriers in this group were examined further. Based on ADMIXTURE analysis, the GP2 EUR LRRK2 p.A419V carriers were found to be highly admixed (Supplementary Table 7a), with 29/35 carriers having relatively lower EUR ancestry (median: 13.86%, range: 4.68% – 70.8%, Supplementary Table 7a) compared to EUR non-carriers (median: 65.75%, range: 4.09% - 82.30 %, Supplementary Table 7b). Notably, 22/35 carriers in the EUR ancestry group were Kazakhstani Central Asian patients. Therefore, these carriers may not be fully representative of the EUR ancestry group. In line with this, linear regression against percentage of genomic admixture in this group indicated that individuals who have less EUR ancestry are more likely to be LRRK2 p.A419V carriers (β = -0.23, SE = 1.78; p = 3.01e-19). In contrast, all the EAS p.A419V carriers demonstrated high EAS ancestry (median: 91.68%, range: 71.9% - 94.1%, Supplementary Table 7c) similar to EAS non-carriers (median: 91.83%, range: 44.24% - 94.91%, Supplementary Table 6d), lending greater confidence to the observed associations within this population.

### Age at onset association analysis in EAS cohorts

Analysis of LRRK2 p.A419V carrier status and AAO in the GP2 EAS PD group (n=73 carriers, AAO 50.3 ± 12.6 years vs. n=2292 non-carriers, AAO 53.4 ± 12.7 years) (Table 3a), revealed an association with an earlier onset of PD by approximately 3 years (β = -3.02 years; SE = 1.49; p = 0.043) after adjusting for sex and 5 PCs (Figure 1). Six LRRK2 p.A419V carriers also carried a concomitant p.G2385R variant (mean AAO: 51.3 ± 13 years), and one patient with young onset, unknown monogenic PD status (AAO: 35 years) was found to have a concomitant p.R1628P variant (Supplementary Table 8). In the replication cohort, which included 103 LRRK2 p.A419V PD carriers and 3,557 non-carriers, linear regression adjusted for sex showed a similarly significant reduction in disease onset by approximately two and a half years (β = -2.79 years, SE = 1.03; p = 0.007). However, after additional adjustment for the first three PCs, the association was no longer statistically significant (β = –0.89 years; SE = 1.01; p = 0.380), despite the directionality of effect remaining consistent, with carriers showing a lower AAO (Table 2b). A fixed-effects meta-analysis combining the discovery and replication cohorts demonstrated a reduced but borderline-significant association (β = -1.55 years, SE = 0.83, p = 0.063; Supplementary Figure 3), supporting a modest effect of the variant on age at onset.

**Figure 1:**
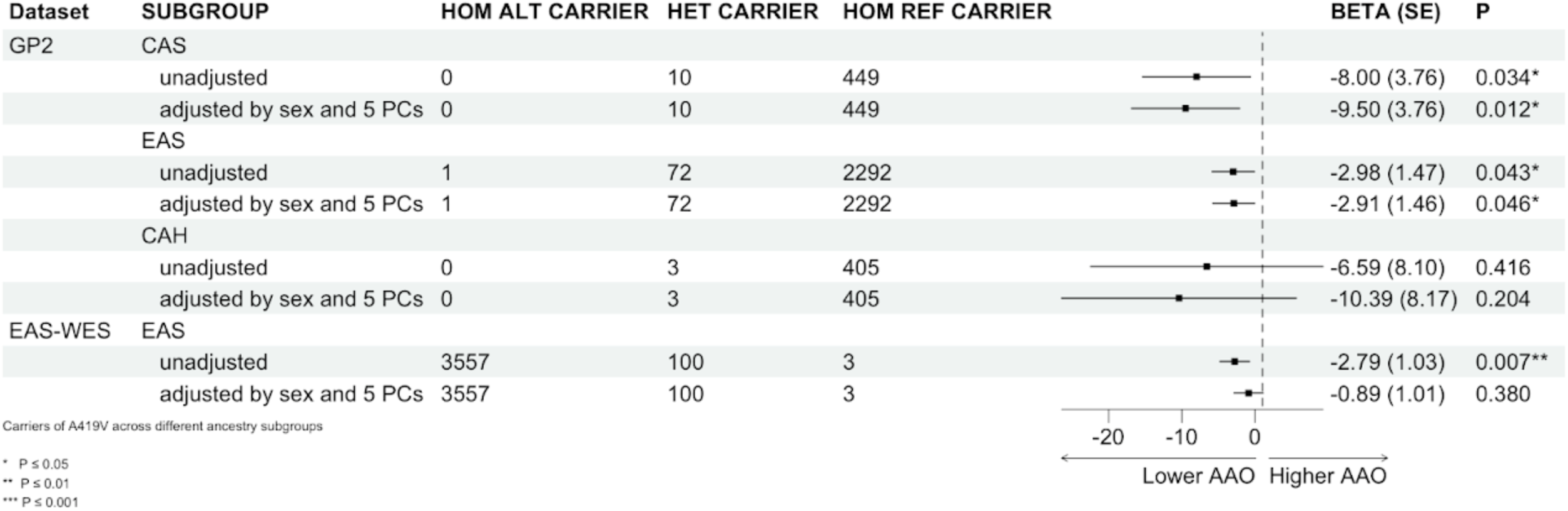
*LRRK2* p.A419V and age at onset (Linear model). Association of *LRRK2* p.A419V with Age of onset were conducted using generalized linear models (GLM) with linear regression under an additive model.

**Table 2:**
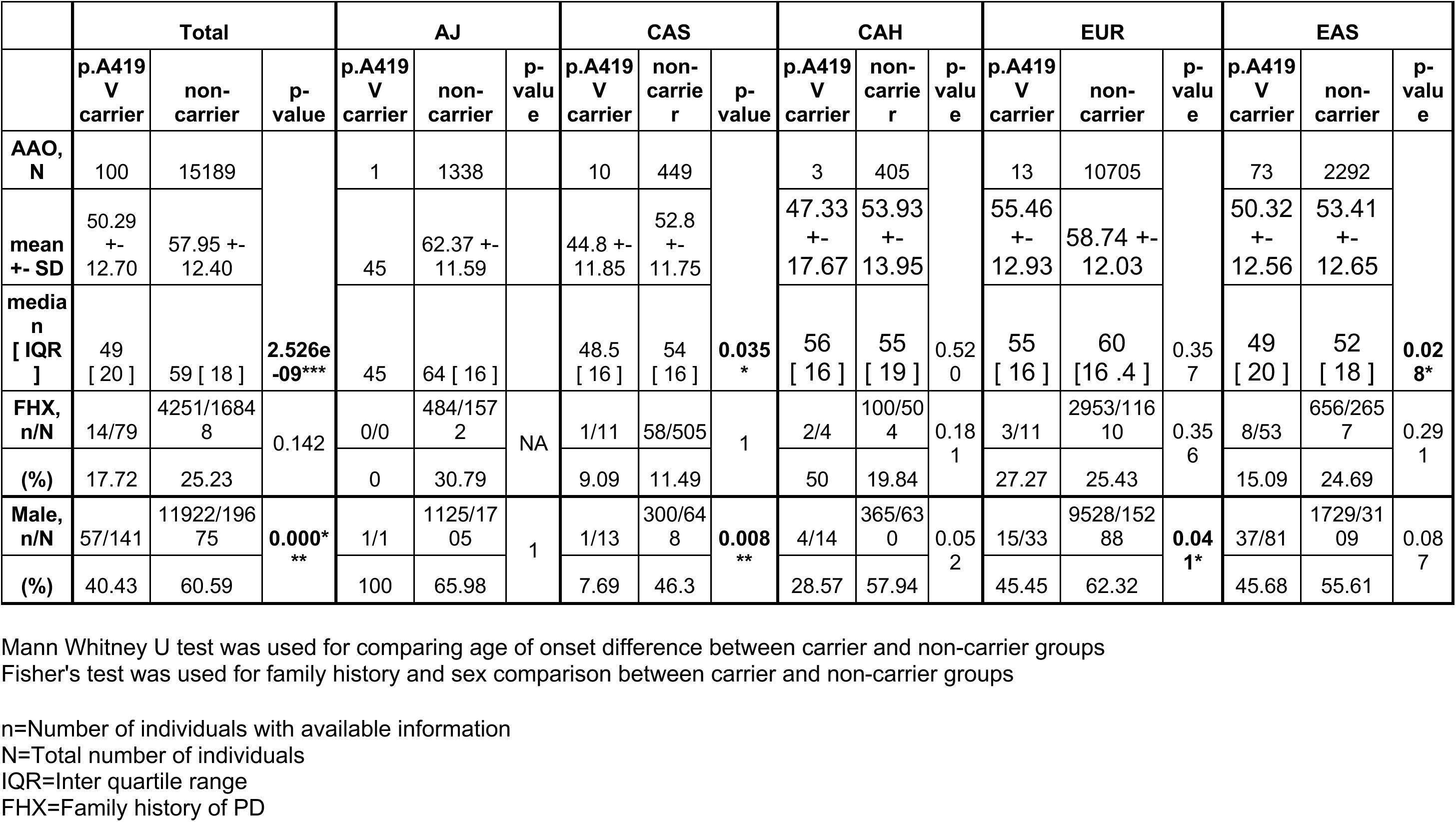
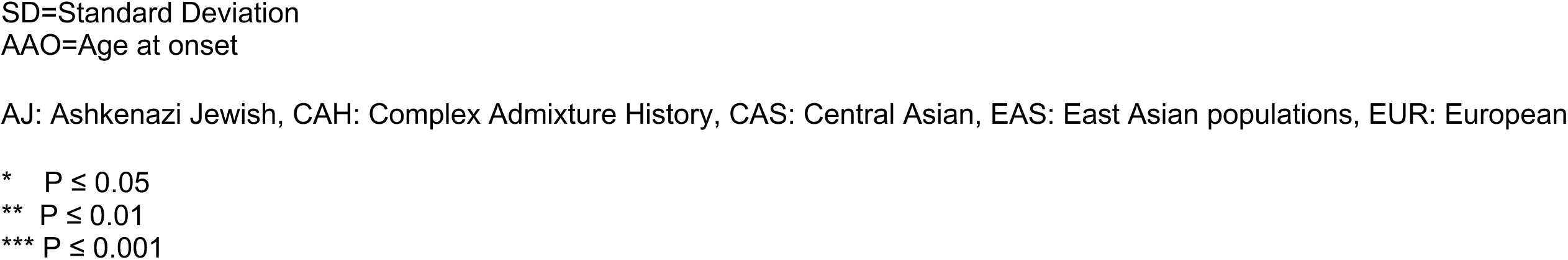
Age at PD onset between *LRRK2* p.A419V PD carriers and non-carriers in the GP2 cohort.

|  | Total |  |  | AJ |  |  | CAS |  |  | CAH |  |  | EUR |  |  | EAS |  |  |
| --- | --- | --- | --- | --- | --- | --- | --- | --- | --- | --- | --- | --- | --- | --- | --- | --- | --- | --- |
|  | p.A419<br>V<br>carrier | non-<br>carrier | p-value | p.A419<br>V<br>carrier | non-<br>carrier | p-<br>valu<br>e | p.A419<br>V<br>carrier | non-<br>carrie<br>r | p-value | p.A419<br>V<br>carrier | non-<br>carrie<br>r | p-<br>valu<br>e | p.A419<br>V<br>carrier | non-<br>carrier | p-<br>valu<br>e | p.A419<br>V<br>carrier | non-<br>carrier | p-<br>valu<br>e |
| **AAO,**<br>**N** | 100 | 15189 | **2.526e-09\*\*\*** | 1 | 1338 |  | 10 | 449 |  | 3 | 405 |  | 13 | 10705 |  | 73 | 2292 |  |
| **mean**<br>**+- SD** | 50.29<br>+-<br>12.70 | 57.95 +-<br>12.40 |  | 45 | 62.37 +-<br>11.59 |  | 44.8 +-<br>11.85 | 52.8<br>+-<br>11.75 |  | 47.33<br>+-<br>17.67 | 53.93<br>+-<br>13.95 |  | 55.46<br>+-<br>12.93 | 58.74 +-<br>12.03 |  | 50.32<br>+-<br>12.56 | 53.41<br>+-<br>12.65 |  |
| **media**<br>**n**<br>**[ IQR**<br>**]** | 49<br>[ 20 ] | 59 [ 18 ] |  | 45 | 64 [ 16 ] |  | 48.5<br>[ 16 ] | 54<br>[ 16 ] | **0.035**<br>**\*** | 56<br>[ 16 ] | 55<br>[ 19 ] | 0.52<br>0 | 55<br>[ 16 ] | 60<br>[16 .4 ] | 0.35<br>7 | 49<br>[ 20 ] | 52<br>[ 18 ] | **0.02**<br>**8\*** |
| **FHX,**<br>**n/N** | 14/79 | 4251/1684<br>8 | 0.142 | 0/0 | 484/157<br>2 | NA | 1/11 | 58/505 | 1 | 2/4 | 100/50<br>4 | 0.18<br>1 | 3/11 | 2953/116<br>10 | 0.35<br>6 | 8/53 | 656/265<br>7 | 0.29<br>1 |
| **(%)** | 17.72 | 25.23 |  | 0 | 30.79 |  | 9.09 | 11.49 |  | 50 | 19.84 |  | 27.27 | 25.43 |  | 15.09 | 24.69 |  |
| **Male,**<br>**n/N** | 57/141 | 11922/196<br>75 | **0.000\***<br>**\*\*** | 1/1 | 1125/17<br>05 | 1 | 1/13 | 300/64<br>8 | **0.008**<br>**\*\*** | 4/14 | 365/63<br>0 | 0.05<br>2 | 15/33 | 9528/152<br>88 | **0.04**<br>**1\*** | 37/81 | 1729/31<br>09 | 0.08<br>7 |
| **(%)** | 40.43 | 60.59 |  | 100 | 65.98 |  | 7.69 | 46.3 |  | 28.57 | 57.94 |  | 45.45 | 62.32 |  | 45.68 | 55.61 |  |

Mann Whitney U test was used for comparing age of onset difference between carrier and non-carrier groups Fisher's test was used for family history and sex comparison between carrier and non-carrier groups
n=Number of individuals with available information N=Total number of individuals IQR=Inter quartile range FHX=Family history of PD
SD=Standard Deviation
AAO=Age at onset
AJ: Ashkenazi Jewish, CAH: Complex Admixture History, CAS: Central Asian, EAS: East Asian populations, EUR: European
\* $P \leq 0.05$
\*\* $P \leq 0.01$
\*\*\* $P \leq 0.001$

The CAS ancestry in the GP2 cohort was observed to show a significant association with earlier AAO (β = -4.25 years; SE = 8.173; p = 0.011). However, given the small number of carriers in this group (n = 10), this should be interpreted cautiously. No significant association between LRRK2 p.A419V and AAO was observed in the CAH group (only present in the GP2 cohort), before or after adjusting for covariates.

Visualization of the predicted LRRK2 structure overlaid with known variants revealed that the p.A419V mutation resides within the ARM domain (amino acid position 12 – 704)^42^ (Figure 2A). Predicted structures of the LRRK2-RAB complexes demonstrated that the binding sites for RAB8A, RAB29, and RAB32 are spatially proximal to the p.A419V mutation site, consistent with experimental observations. Notably, the predicted binding site for RAB10 deviates from the ARM domain, despite experimental evidence suggesting similar binding motifs with other Rab proteins^43,44^.

**Figure 2.**
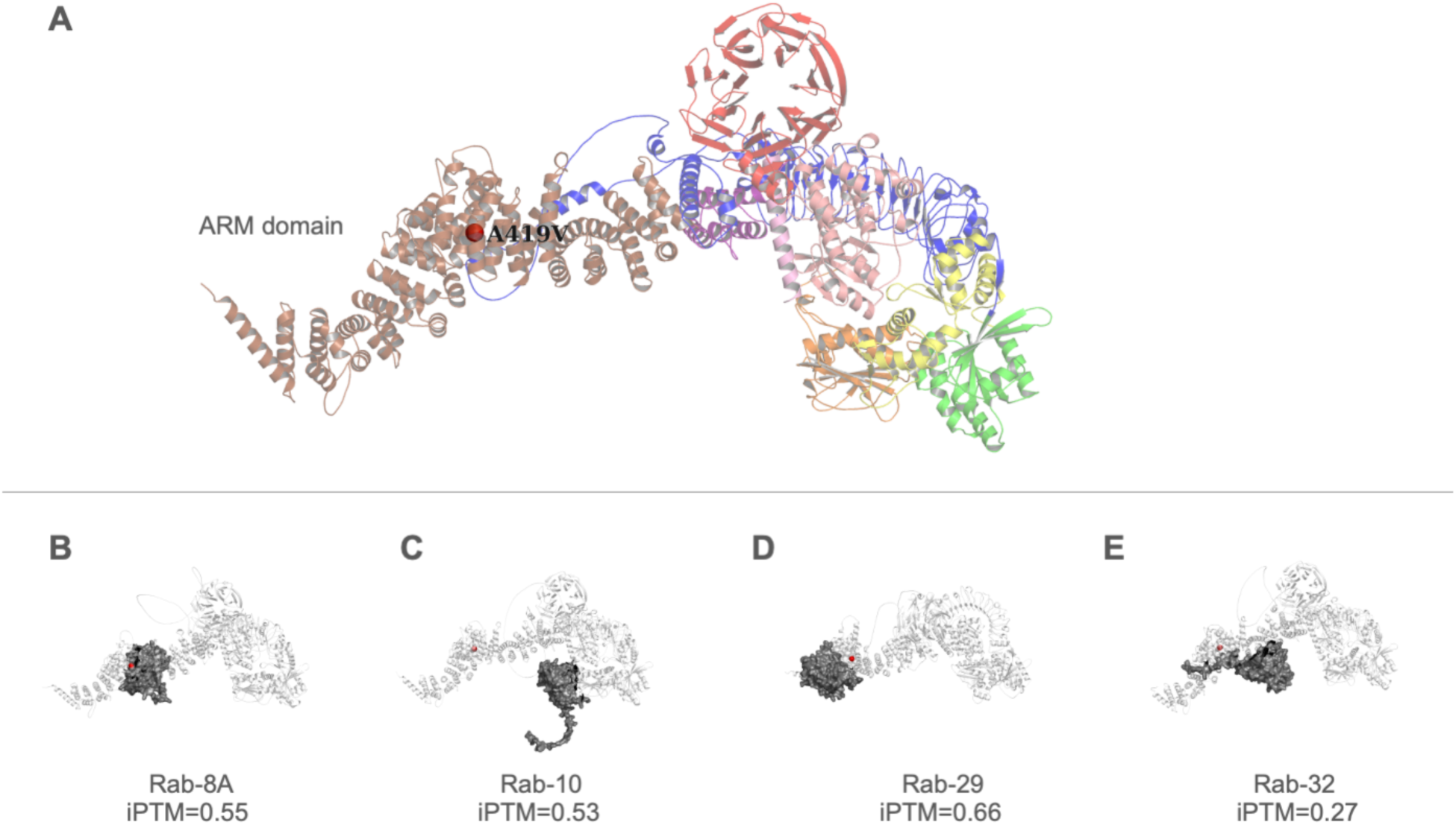
Mutation sites and predicted Rab binding of LRRK2. Visualization of the predicted LRRK2 structure with different coloured domains (armadillo repeat (ARM) domain in brown/tan, ankyrin repeat (ANK) domain in pink, leucine-rich repeat (LRR) domain in blue, ROC (Ras of complex proteins) GTPase domain in yellow, COR (C-terminal of ROC) domain in orange, kinase domain in green, and the WD40 domain in red). The alanine residue at position 419, substituted by valine in the p.A419V variant, is indicated by a red sphere within the ARM domain.

## Discussion

We investigated the genetic association between the LRRK2 p.A419V variant and PD risk and AAO, leveraging data across several cohorts, including the multi-ancestry GP2, AMP-PD, UKB, AOU cohorts, an EAS exome dataset and a Japanese dataset, totaling 40,287 PD cases and 168,681 controls. This study resolves inconsistencies in previous reports by providing robust evidence that the LRRK2 p.A419V is significantly associated with PD in both the EAS and EUR populations, with an approximate OR of 2.0. This is comparable with other LRRK2 risk variants in EAS populations, p.G2385R (OR ∼ 2.3)^45^ and p.R1628P (OR ∼ 1.8)^46^.

The variant is rare in EUR (MAF = 0.03%) and even rarer or absent in these AAC, AFR, AJ, AMR, FIN, and MDE populations’ samples. Compared to the other LRRK2 risk variants p.R1628P and p.G2385R (MAFs between 5-10% in cases and 2-5% in controls) (Lim et al. 2019), in the overall EAS cohorts in this study, the p.A419V variant is much less common (MAFTotal = 0.91%, MAFcases = 1.35% and MAFcontrols = 0.54%). Interestingly, this variant was also seen in other Asian populations, such as CAS (MAFTotal = 0.71%, MAFcases = 0.97% and MAFcontrols = 0.38%) and SAS (MAFTotal = 0.07%, MAFcases = 0.12% and MAFcontrols = 0.05%), compared to non-Asian populations, suggesting this variant may be more relevant to individuals of ‘East-Central-South’ Asian ancestries. The MAF seen in the SAS population in this study (0.07%) compares to 0.4% in a SAS GWAS by Kishore et al. 2025^16^ (n=11,170), with gnomAD reporting an MAF of 0.02%. Thus far, the p.A419V variant has not been investigated as a risk variant in SAS populations. In CAS populations, a previous study found no significant association between the p.A419V variant and PD^15^, and this has also been observed in an unpublished extended sample set of 655 cases (MAF = 1.38%) and 528 controls (MAF = 0.85%) (personal communication, RK).

Only one LRRK2 p.A419V haplotype (Haplotype 9) was seen in both cases and controls, but association analysis for this haplotype is not possible due to its low frequency in both cases and controls. No disease-specific haplotype enrichment was observed. Haplotype analysis was not performed in the EAS-WES cohort due to limited variants from exome data.

Regarding the heritability of AAO concerning LRRK2 risk variants, a directionality towards an earlier AAO has been reported. Specifically, the combination of the p.G2385R-p.R1628P-p.S1647T variants was found responsible for lowering the AAO by approximately 8 years^30^. Regarding the LRRK2 p.A419V variant, a previous EAS study of 2,685 cases from mainland China did not find any association with an earlier AAO^31^. However, our study showed a modest effect on AAO in the GP2 EAS and EAS exome cohorts. We found LRRK2 p.A419V carriers developing PD approximately 3 years earlier than non-carriers, although we note the lack of robust association. A limitation of interpreting this result is the analysis of a single gene variant. Previous GWAS on AAO in European populations have identified variants in the TMEM175 and SNCA genes^47^ and BST1^48^. Only two studies on AAO in the East Asian PD population were conducted. One was a meta-analysis of GWAS, which highlighted a SNP (rs9783733) in the novel NDN; PWRN4 locus, delaying AAO by 2.43 years, most significantly in male patients, as well as a suggestive signal in SNCA (rs3775458), which lowers AAO by 1.36 years^49^. In the other and more recent study, SNPs in LD with ALCAM were associated with an earlier AAO by 3.47 years in a Korean AAO-GWAS^50^. Additionally, polygenic risk scores (PRS) across multiple studies have shown a strong inverse correlation with AAO^49–54^. Thus, the contribution of the LRRK2 p.A419V variant on AAO will need to be evaluated more deeply in the context of other genome-wide genetic modifiers.

While an association with LRRK2 p.A419V was initially found in the GP2 EUR group, closer inspection of these carriers revealed that 22/35 of these patients were of Central Asian origin. As this variant is very rare in Europeans and in Central Europeans^55^ and its association with PD has not been reported in individuals of EUR ancestry despite multiple GWAS and LRRK2 studies^4,14,21,56^, we postulate that the association in our study may be an artefact due to inheritance of the LRRK2 p.A419V variant from the EAS/CAS chromosomal regions in these individuals. Resolution of admixture in the EUR-CAS individuals could be performed through local ancestry analysis. However, this was not currently possible as the number of control CAS genomes was insufficient. The issue of limited control individuals is not uncommon in underrepresented populations^57^ and presently poses a challenge in interpreting the contribution of rare variants such as p.A419V. Large-scale initiatives such as GP2, which is sequencing both PD cases and ancestry-matched controls, and existing or emerging national genome projects of several underrepresented populations contribute to bridging this gap. These include Kazakhstan (Central Asia)^58–60^, China (East Asia)^61^, Singapore (Southeast Asia)^62^, and the Americas^6,63^. Future efforts will play a crucial role in providing a more accurate interpretation of disease-relevant genomic variants.

The alanine residue at position 419 in LRRK2 is highly conserved (ConSurf score 9/9), suggesting that mutations at this site may significantly alter protein function. The p.A419V variant lies within the LRRK2 armadillo repeat domain (ARM), which mediates the interaction between LRRK2 and multiple Rab GTPases^43,64,65^. Thus, facilitating recruitment of LRRK2 to intracellular vesicle trafficking pathways, including the trans-Golgi network and endolysosomal system^66–69^. Recent studies indicate that the variant is located in a distinct region in the ARM domain, termed site #1 (encompassing amino acids 360-450), which is reported to be the precise binding site of RAB29, RAB8A, and RAB10^43,44^. Together, the Rab substrates anchor LRRK2 to membranes, enabling it to acquire a more active state through enhanced GTPase phosphorylation, in a proposed ‘feed-forward pathway’ ^44,70^. Cells carrying variants neighbouring p.A149V, p.L403A, and p.K439E interfere with optimal binding of Rab8A and Rab29 substrates^44^ and recently the p.[L119P;L488P] variants were also shown to affect the binding affinity of RAB8A^71^, and it could be postulated that the p.A419V variant may have similar effects. However, this has yet to be demonstrated and would be best addressed with in vitro and in vivo models.

Through LRRK2 protein modelling with Alphafold3, the predicted model with the available regions of the X-ray structure as published in Myasnikov et al., 2021^42^, yielded acceptable agreement (RMSD = 0.992), underscoring the value of the predicted model for visualizing the ARM domain. The predicted interaction interfaces between LRRK2 and RAB proteins placed the p.A419V variant in close proximity to the binding regions for RAB8A, RAB29, and RAB32, which tallies with experimental data^43,44^. However, while these predictions offer valuable insights, they are inherently limited in defining the precise binding sites. The deviation observed for RAB10, whose predicted site lies outside of the ARM domain, may arise due to the AlphaFold3 model itself and/or the possibility that LRRK2 exists as a dimer during RAB protein interactions. Although these limitations exist, the predicted structures are useful for visualization and will guide future structural and functional investigations, particularly as current X-ray structures lack the ARM domain. This finding suggests if the p.A419V variant causes conformational changes to LRRK2, this may have an effect on binding affinity to RAB proteins through the ARM domain, and this may lead to disrupted LRRK2-RAB signaling.

In conclusion, the LRRK2 p.A419V variant is a rare but significant risk factor for PD in EAS individuals. Further research on this variant, along with LRRK2 p.R1628P and p.G2385R, is needed to gain a more comprehensive understanding of the genetic architecture of EAS PD populations. In the emerging era of LRRK2-directed treatments, this study highlights that variants in the ARM domain, the site of several RAB substrate binding, may be a rare but relevant target for further clinical, biomarker, and therapeutic studies.

## Acknowledgements

We gratefully acknowledge all the participants for their contributions, without whom this research would not have been possible. Data and code used to prepare this article were obtained from the Global Parkinson’s Genetics Program (GP2). GP2 is funded by the Aligning Science Across Parkinson’s (ASAP) initiative and implemented by The Michael J. Fox Foundation for Parkinson’s Research (https://gp2.org). For a complete list of GP2 members, see https://gp2.org. Data used to prepare this article were obtained from the Accelerating Medicines Partnership® (AMP®) Parkinson’s Disease (AMP® PD) Knowledge Platform. For up-to-date information on the study, visit https://www.amp-pd.org. ACCELERATING MEDICINES PARTNERSHIP and AMP are registered service marks of the US Department of Health and Human Services. We thank the research participants for consenting to this study. Additionally, data used in this article were obtained from the UK Biobank, the All of Us Research Program, an East Asian whole-exome dataset, and a Japanese whole-exome dataset. This research has been conducted using the UK Biobank Resource under Application Number 33601.

The All of Us Research Program is supported by the National Institutes of Health, Office of the Director: Regional Medical Centers: 1 OT2 OD026549; 1 OT2 OD026554; 1 OT2 OD026557; 1 OT2 OD026556; 1 OT2 OD026550; 1 OT2 OD 026552; 1 OT2 OD026553; 1 OT2 OD026548; 1 OT2 OD026551; 1 OT2 OD026555; IAA #: AOD 16037; Federally Qualified Health Centers: HHSN 263201600085U; Data and Research Center: 5 U2C OD023196; Biobank: 1 U24 OD023121; The Participant Center: U24 OD023176; Participant Technology Systems Center: 1 U24 OD023163; Communications and Engagement: 3 OT2 OD023205; 3 OT2 OD023206; and Community Partners: 1 OT2 OD025277; 3 OT2 OD025315; 1 OT2 OD025337; 1 OT2 OD025276. In addition, the All of Us Research Program would not be possible without the partnership of its participants.

This research was supported [in part] by the Intramural Research Program of the National Institutes of Health (NIH). The contributions of the NIH author(s) are considered Works of the United States Government. The findings and conclusions presented in this paper are those of the author(s) and do not necessarily reflect the views of the NIH or the U.S. Department of Health and Human Services.

## Authors contributions

AAA, AHT, and LKS conceptualized the manuscript. AAA, LKS, SBC, MTP, EGYC, RK conducted the formal analysis. AAA, LKS, SBC, and MTP wrote the first draft of the manuscript. All authors reviewed, edited, and approved the final version of the manuscript for submission.

## Statements and declarations

## Ethical considerations

SingHealth Centralized Institutional Review Board (CIRB 2002/008/A and 2019/2334) and Nanyang Technological University Institutional Review Board (IRB-2016-08-011)

Ethics committee of Juntendo University, Tokyo, Japan, and all participants provided written informed consent to participate in the research described in this study (M08&#8208;0477&#8208;M09)

International Genetics Collaboration (IGC) CI: Prof H Houlden Sponsor EDGE ID: 146653 REC Ref: IRAS: 310045 Protocol V1.1 22/06/2022

## Data availability

All GP2 data is hosted in collaboration with the Accelerating Medicines Partnership in Parkinson’s disease, and is available via application on the website (https://amp-pd.org/register-for-amp-pd;https://doi.org/10.5281/zenodo.7904832). Genotyping imputation, quality control, ancestry prediction, and processing was performed using GenoTools v1.0, publicly available on GitHub (https://github.com/GP2code/GenoTools). https://github.com/AAA-lab-genetic/Multiancestry_LRRK2_p.A419V)

## Supplementary Materials

Supplementary Table: Supplementary table

Supplementary Figure: Supplementary figure

## References

1 Ross, O. A. et al. Association of LRRK2 exonic variants with susceptibility to Parkinson’s disease: a case-control study. Lancet Neurol 10, 898–908 (2011). 10.1016/s1474-4422(11)70175-2

2 Krüger, C. et al. Updated MDSGene review on the clinical and genetic spectrum of LRRK2 variants in Parkinsońs disease. NPJ Parkinsons Dis 11, 30 (2025). 10.1038/s41531-025-00881-9

3 Lim, S.-Y. et al. Clinical and functional evidence for the pathogenicity of the LRRK2 p.Arg1067Gln variant. npj Parkinson’s Disease 11, 34 (2025). 10.1038/s41531-025-00884-6

4 Program, T. G. P. s. G. & Leonard, H. L. Novel Parkinson’s Disease Genetic Risk Factors Within and Across European Populations. medRxiv, 2025.2003.2014.24319455 (2025). 10.1101/2025.03.14.24319455

5 Foo, J. N. et al. Identification of Risk Loci for Parkinson Disease in Asians and Comparison of Risk Between Asians and Europeans: A Genome-Wide Association Study. JAMA Neurol 77, 746–754 (2020). 10.1001/jamaneurol.2020.0428

6 Lim, S. Y. et al. Uncovering the genetic basis of Parkinson’s disease globally: from discoveries to the clinic. Lancet Neurol 23, 1267–1280 (2024). 10.1016/s1474-4422(24)00378-8

7 Di Fonzo, A. et al. Comprehensive analysis of the LRRK2 gene in sixty families with Parkinson’s disease. Eur J Hum Genet 14, 322–331 (2006). 10.1038/sj.ejhg.5201539

8 Tan, E. K. et al. Multiple LRRK2 variants modulate risk of Parkinson disease: a Chinese multicenter study. Hum Mutat 31, 561–568 (2010). 10.1002/humu.21225

9 Wu, X. et al. Quantitative assessment of the effect of LRRK2 exonic variants on the risk of Parkinson’s disease: a meta-analysis. Parkinsonism Relat Disord 18, 722–730 (2012). 10.1016/j.parkreldis.2012.04.013

10 Heckman, M. G. et al. Population-specific frequencies for LRRK2 susceptibility variants in the Genetic Epidemiology of Parkinson’s Disease (GEO-PD) Consortium. Mov Disord 28, 1740–1744 (2013). 10.1002/mds.25600

11 Li, K. et al. LRRK2 A419V variant is a risk factor for Parkinson’s disease in Asian population. Neurobiol Aging 36, 2908.e2911–2905 (2015). 10.1016/j.neurobiolaging.2015.07.012

12 Gopalai, A. A. et al. Lack of association between the LRRK2 A419V variant and Asian Parkinson’s disease. Ann Acad Med Singap 42, 237–240 (2013).

13 Pan, H. et al. Genome-wide association study using whole-genome sequencing identifies risk loci for Parkinson’s disease in Chinese population. npj Parkinson’s Disease 9, 22 (2023). 10.1038/s41531-023-00456-6

14 Kim, J. J. et al. Multi-ancestry genome-wide association meta-analysis of Parkinson’s disease. Nat Genet 56, 27–36 (2024). 10.1038/s41588-023-01584-8

15 Kaiyrzhanov, R. et al. LRRK2 Mutations and Asian Disease-Associated Variants in the First Parkinson’s Disease Cohort from Kazakhstan. Parkinsons Dis 2020, 2763838 (2020). 10.1155/2020/2763838

16 Kishore, A. et al. Deciphering the Genetic Architecture of Parkinson’s Disease in India. medRxiv, 2025.2002.2017.25322132 (2025). 10.1101/2025.02.17.25322132

17 Do, M. D. et al. Clinical and genetic analysis of Vietnamese patients diagnosed with early-onset Parkinson’s disease. Brain Behav 13, e2950 (2023). 10.1002/brb3.2950

18 Kanaya, Y. et al. Analysis of genetic risk factors in Japanese patients with Parkinson’s disease. J Hum Genet 66, 957–964 (2021). 10.1038/s10038-021-00910-4

19 Thanprasertsuk, S. et al. Levodopa-induced dyskinesia in early-onset Parkinson’s disease (EOPD) associates with glucocerebrosidase mutation: A next-generation sequencing study in EOPD patients in Thailand. PLoS One 18, e0293516 (2023). 10.1371/journal.pone.0293516

20 Park, K. W. et al. Ethnicity- and sex-specific genome wide association study on Parkinson’s disease. NPJ Parkinsons Dis 9, 141 (2023). 10.1038/s41531-023-00580-3

21 Nalls, M. A. et al. Identification of novel risk loci, causal insights, and heritable risk for Parkinson’s disease: a meta-analysis of genome-wide association studies. Lancet Neurol 18, 1091–1102 (2019). 10.1016/s1474-4422(19)30320-5

22 Rizig, M. et al. Identification of genetic risk loci and causal insights associated with Parkinson’s disease in African and African admixed populations: a genome-wide association study. Lancet Neurol 22, 1015–1025 (2023). 10.1016/s1474-4422(23)00283-1

23 Cornejo-Olivas, M. et al. Variable frequency of LRRK2 variants in the Latin American research consortium on the genetics of Parkinson’s disease (LARGE-PD), a case of ancestry. npj Parkinson’s Disease 3, 19 (2017). 10.1038/s41531-017-0020-6

24 William, M. B. et al. The p.Gly2019Ser is a common LRRK2 pathogenic variant among Egyptians with familial and sporadic Parkinson’s disease. npj Parkinson’s Disease 10, 215 (2024). 10.1038/s41531-024-00826-8

25 Kalogeropulou, A. F. et al. Impact of 100 LRRK2 variants linked to Parkinson’s disease on kinase activity and microtubule binding. Biochem J 479, 1759–1783 (2022). 10.1042/bcj20220161

26 Toh, T. S., Lit, Lei Cheng Lit, Lim, Shen-Yang, Hor, Jia Wei, Lew, Cindy Choey Yee, Khairul Anuar, Anis Nadhirah, Tay, Yi Wen, Lim, Jia Lun, Ahmad-Annuar Azlina, Tan, Eng King, Alessi, Dario R., Sammler, Esther, Tan, Ai Huey. in MDS 2025 Abstract (2025).

27 Lange, L. M. et al. Elucidating causative gene variants in hereditary Parkinson’s disease in the Global Parkinson’s Genetics Program (GP2). NPJ Parkinsons Dis 9, 100 (2023). 10.1038/s41531-023-00526-9

28 Towns, C. et al. Defining the causes of sporadic Parkinson’s disease in the global Parkinson’s genetics program (GP2). npj Parkinson’s Disease 9, 131 (2023). 10.1038/s41531-023-00533-w

29 Chew, E. G. Y. et al. Exome sequencing in Asian populations identifies low-frequency and rare coding variation influencing Parkinson’s disease risk. Nature Aging 5, 205–218 (2025). 10.1038/s43587-024-00760-7

30 Xiao, B. et al. Association of LRRK2 Haplotype With Age at Onset in Parkinson Disease. JAMA Neurol 75, 127–128 (2018). 10.1001/jamaneurol.2017.3363

31 Song, T. et al. Clinical features and progression of Parkinson’s disease with LRRK2 variants: A prospective study. Ann Clin Transl Neurol 12, 34–42 (2025). 10.1002/acn3.52244

32 Tadaka, S. et al. jMorp: Japanese Multi-Omics Reference Panel update report 2023. Nucleic Acids Res 52, D622–d632 (2024). 10.1093/nar/gkad978

33 Vitale, D. et al. GenoTools: an open-source Python package for efficient genotype data quality control and analysis. G3 (Bethesda) 15 (2025). 10.1093/g3journal/jkae268

34 Alexander, D. H., Novembre, J. & Lange, K. Fast model-based estimation of ancestry in unrelated individuals. Genome Res 19, 1655–1664 (2009). 10.1101/gr.094052.109

35 Bandres-Ciga, S. et al. NeuroBooster Array: A Genome-Wide Genotyping Platform to Study Neurological Disorders Across Diverse Populations. Mov Disord 39, 2039–2048 (2024). 10.1002/mds.29902

36 Brolin, K. et al. Insights on Genetic and Environmental Factors in Parkinson’s Disease from a Regional Swedish Case-Control Cohort. J Parkinsons Dis 12, 153–171 (2022). 10.3233/jpd-212818

37 Balduzzi, S., Rücker, G. & Schwarzer, G. How to perform a meta-analysis with R: a practical tutorial. Evid Based Ment Health 22, 153–160 (2019). 10.1136/ebmental-2019-300117

38 Satake, W. et al. Genome-wide association study identifies common variants at four loci as genetic risk factors for Parkinson’s disease. Nat Genet 41, 1303–1307 (2009). 10.1038/ng.485

39 Foo, J. N. et al. Genome-wide association study of Parkinson’s disease in East Asians. Hum Mol Genet 26, 226–232 (2017). 10.1093/hmg/ddw379

40 Abramson, J. et al. Accurate structure prediction of biomolecular interactions with AlphaFold 3. Nature 630, 493–500 (2024). 10.1038/s41586-024-07487-w

41 UniProt: the Universal Protein Knowledgebase in 2023. Nucleic Acids Res 51, D523–d531 (2023). 10.1093/nar/gkac1052

42 Myasnikov, A. et al. Structural analysis of the full-length human LRRK2. Cell 184, 3519–3527.e3510 (2021). 10.1016/j.cell.2021.05.004

43 McGrath, E., Waschbüsch, D., Baker, B. M. & Khan, A. R. LRRK2 binds to the Rab32 subfamily in a GTP-dependent manner via its armadillo domain. Small GTPases 12, 133–146 (2021). 10.1080/21541248.2019.1666623

44 Vides, E. G. et al. A feed-forward pathway drives LRRK2 kinase membrane recruitment and activation. Elife 11 (2022). 10.7554/eLife.79771

45 Shu, L., Zhang, Y., Sun, Q., Pan, H. & Tang, B. A Comprehensive Analysis of Population Differences in LRRK2 Variant Distribution in Parkinson’s Disease. Front Aging Neurosci 11, 13 (2019). 10.3389/fnagi.2019.00013

46 Zhang, Y. et al. Genetic Analysis of LRRK2 R1628P in Parkinson’s Disease in Asian Populations. Parkinsons Dis 2017, 8093124 (2017). 10.1155/2017/8093124

47 Blauwendraat, C. et al. Parkinson’s disease age at onset genome-wide association study: Defining heritability, genetic loci, and α-synuclein mechanisms. Mov Disord 34, 866–875 (2019). 10.1002/mds.27659

48 Grover, S. et al. Genome-wide Association and Meta-analysis of Age at Onset in Parkinson Disease: Evidence From the COURAGE-PD Consortium. Neurology 99, e698–e710 (2022). 10.1212/wnl.0000000000200699

49 Li, C. et al. Genetic Modifiers of Age at Onset for Parkinson’s Disease in Asians: A Genome-Wide Association Study. Movement Disorders 36, 2077–2084 (2021). 10.1002/mds.28621

50 Hwang, Y. S. et al. Identification of Novel Genetic Loci Affecting Age at Onset of Parkinson’s Disease: A Genome-wide Association Study. Movement Disorders 40, 77–86 (2025). 10.1002/mds.30047

51 Escott-Price, V. et al. Common polygenic variation enhances risk prediction for Alzheimer’s disease. Brain 138, 3673–3684 (2015). 10.1093/brain/awv268

52 Pavelka, L. et al. Age at onset as stratifier in idiopathic Parkinson’s disease – effect of ageing and polygenic risk score on clinical phenotypes. npj Parkinson’s Disease 8, 102 (2022). 10.1038/s41531-022-00342-7

53 Huang, Y. et al. Risk factors associated with age at onset of Parkinson’s disease in the UK Biobank. npj Parkinson’s Disease 10, 3 (2024). 10.1038/s41531-023-00623-9

54 Gabbert, C. et al. The combined effect of lifestyle factors and polygenic scores on age at onset in Parkinson’s disease. Scientific Reports 14, 14670 (2024). 10.1038/s41598-024-65640-x

55 Skorvanek, M. et al. LRRK2 mutations in Parkinson’s disease patients from Central Europe: A case control study. Parkinsonism Relat Disord 83, 110–112 (2021). 10.1016/j.parkreldis.2020.12.021

56 Simpson, C. et al. Prevalence of ten LRRK2 variants in Parkinson’s disease: A comprehensive review. Parkinsonism Relat Disord 98, 103–113 (2022). 10.1016/j.parkreldis.2022.05.012

57 Schumacher-Schuh, A. F. et al. Underrepresented Populations in Parkinson’s Genetics Research: Current Landscape and Future Directions. Mov Disord 37, 1593–1604 (2022). 10.1002/mds.29126

58 Akilzhanova, A. et al. The First Kazakh Whole Genomes: The First Report of NGS Data. Cent Asian J Glob Health 3, 146 (2014). 10.5195/cajgh.2014.146

59 Kairov, U. et al. Whole-genome sequencing data of Kazakh individuals. BMC Research Notes 14, 45 (2021). 10.1186/s13104-021-05464-4

60 The GenomeAsia 100K Project enables genetic discoveries across Asia. Nature 576, 106–111 (2019). 10.1038/s41586-019-1793-z

61 Choi, J. et al. A whole-genome reference panel of 14,393 individuals for East Asian populations accelerates discovery of rare functional variants. Science Advances 9, eadg6319 (2023). doi:10.1126/sciadv.adg6319

62 Wu, D. et al. Large-Scale Whole-Genome Sequencing of Three Diverse Asian Populations in Singapore. Cell 179, 736–749.e715 (2019). 10.1016/j.cell.2019.09.019

63 Borda, V. et al. Genetics of Latin American Diversity Project: Insights into population genetics and association studies in admixed groups in the Americas. Cell Genom 4, 100692 (2024). 10.1016/j.xgen.2024.100692

64 Zhu, H. et al. Rab29-dependent asymmetrical activation of leucine-rich repeat kinase 2. Science 382, 1404–1411 (2023). 10.1126/science.adi9926

65 Taylor, M. & Alessi, D. R. Advances in elucidating the function of leucine-rich repeat protein kinase-2 in normal cells and Parkinson’s disease. Current Opinion in Cell Biology 63, 102–113 (2020). 10.1016/j.ceb.2020.01.001

66 Bonet-Ponce, L. & Cookson, M. R. LRRK2 recruitment, activity, and function in organelles. Febs j 289, 6871–6890 (2022). 10.1111/febs.16099

67 Liu, Z. et al. LRRK2 phosphorylates membrane-bound Rabs and is activated by GTP-bound Rab7L1 to promote recruitment to the trans-Golgi network. Hum Mol Genet 27, 385–395 (2018). 10.1093/hmg/ddx410

68 Purlyte, E. et al. Rab29 activation of the Parkinson’s disease-associated LRRK2 kinase. Embo j 37, 1–18 (2018). 10.15252/embj.201798099

69 Steger, M. et al. Phosphoproteomics reveals that Parkinson’s disease kinase LRRK2 regulates a subset of Rab GTPases. Elife 5 (2016). 10.7554/eLife.12813

70 Alessi, D. R. & Pfeffer, S. R. Leucine-Rich Repeat Kinases. Annu Rev Biochem 93, 261–287 (2024). 10.1146/annurev-biochem-030122-051144

71 Vela-Desojo, L. et al. A new LRRK2 variant in a family with Parkinson’s disease affects binding to RAB8A. npj Parkinson’s Disease 11, 154 (2025). 10.1038/s41531-025-00989-y

